# Gender- and Age-Specific Associations of Visit-To-Visit Blood Pressure Variability with Incident Generalized Anxiety Disorder

**DOI:** 10.1101/2020.12.26.20248868

**Authors:** Jiandong Zhou, Sharen Lee, Wing Tak Wong, Keith Sai Kit Leung, Ronald Hang Kin Lam, Prudence Shun Hay Leung, Tong Liu, Carlin Chang, Bernard Man Yung Cheung, Gary Tse, Qingpeng Zhang

## Abstract

**Background:** There is a bidirectional relationship between blood pressure variability (BPV) and generalized anxiety disorder (GAD), but few studies have examined the gender- and age-specific effects of visit-to-visit BPV on GAD incidence. We examined the predictive value of BPV for the incidence of GAD in a family clinic cohort.

**Methods:** Consecutive patients with a first attendance to family medicine clinics in Hong Kong between January 1^st^, 2000, and December 31^st^, 2002, with at least three blood pressure measurements available thereafter were included. The primary endpoint was incident GAD as identified by ICD-9 coding from the local Clinical Data Analysis and Reporting System.

**Results:** This study included 48023 (50% males) patients with a median follow-up of 224 (IQR: 217-229) months. Females were more likely to develop GAD compared to males (incidence rate: 7% vs. 2%), as were patients of older age. Significant univariate predictors were female gender, older age, pre-existing cardiovascular diseases, respiratory diseases, diabetes mellitus, hypertension, and gastrointestinal diseases, various laboratory examinations and the number of blood pressure measurements. Higher baseline, maximum, minimum, SD, CV, and variability score of diastolic blood pressure significantly predicted GAD, as did all systolic blood pressure measures (baseline, latest, maximum, minimum, mean, median, variance, SD, RMS, CV, variability score).

**Conclusions:** The relationships between longer term visit-to-visit BPV and incident GAD were identified. Female and older patients with higher blood pressure and higher BPV were at the highest risks of GAD.

## Introduction

Generalized anxiety disorder (GAD) is a common group of conditions characterized by excessive worry associated with fatigue, restlessness, muscle tension, irritability, sleeping difficulty, and concentration problems. It is a major public health problem in many countries, damaging not only psychological health, but also physical health and quality of life. There is a bidirectional relationship between blood pressure variability (BPV) and GAD. GAD can exert effects on BPV. Patients with depressive symptoms presented a significantly lower night-time systolic BP fall than non-depressed patients after controlling for age, sex, and traditional cardiovascular risk factors (Scuteri et al., 2009). The control of negative emotions has been shown to influence BP control and BPV (Symonides et al., 2014). Conversely, increased beat-to-beat BPV has been associated with incident GAD (Virtanen et al., 2003). Longer term visit-to-visit BPV has also been reported as an independent predictor of cognitive impairment in several cohort studies (Nagai, Hoshide, Ishikawa, Shimada, & Kario, 2012; Sabayan et al., 2013; Yano et al., 2014). With the widespread measurement of BP measurements at home, fluctuations in blood pressure, as well as very high or low BP readings at home can cause anxiety in patients. However, few previous studies have examined the longitudinal relationship between BPV and anxiety disorders in older cohorts. In this study, we investigated the gender- and age-specific associations of longer period visit-to-visit BPV with the incidence of GAD.

## Methods

### Research design and data sources

The study was approved by The Joint Chinese University of Hong Kong – New Territories East Cluster Clinical Research Ethics Committee and Institutional Review Board of the University of Hong Kong/Hospital Authority Hong Kong West Cluster. This was a retrospective cohort study of family medicine patients who attended family medicine clinics between 1^st^ January 2000 and 31^st^ March 2002 in the Hong Kong public sector. Patients with at least three BP measurements before being diagnosed with GAD were included to calculate the variability measures. The patients were identified from the Clinical Data Analysis and Reporting System (CDARS), a territory-wide database that centralizes patient information from individual local hospitals to establish comprehensive medical data, including clinical characteristics, disease diagnosis, laboratory results, and medication prescription details. The system has been previously used by both our team and other teams in Hong Kong (Ju et al., 2020; Lee et al., 2020). Data were obtained regarding consecutive patients diagnosed with GAD, excluding those who died or discharged within 24 hours after the first diastolic/systolic blood pressure measurement and those with fewer than three diastolic/systolic blood pressure measurements (study baseline). Mortality data were obtained from the Hong Kong Death Registry, a population-based official government registry with the registered death records of all Hong Kong citizens. Data on the clinical characteristics, disease diagnosis, laboratory results (including complete blood count, renal and liver function tests, glycemic and lipid profile, diastolic/systolic BP), and medication prescription details were extracted. Patients with GAD were identified with the diagnosis codes 311, 296.3, 296.2, 308, 300.4, 292.84, 298, 300.02, 291.89, 293.84, 292.89, 294.9, 300.2, 309.24, 300.01, 309.21. The ICD-9 codes for past comorbidities and historical medication prescriptions are detailed in the **Supplementary Appendix**.

### Primary outcome and statistical analysis

The primary outcome was incident cases of GAD from the study baseline in a time-to-event analysis. Follow-up was carried out until 31^st^ December 2019. We extracted the baseline/latest/maximum/minimum values of diastolic and systolic blood pressure, and calculated the temporal variability measures of diastolic and systolic BP: 1) mean, 2) median, 3) standard deviation (SD), 4) root mean square (RMS) by first squaring all blood pressure values then performing square root of the mean of the squares, 5) coefficient of variation (CV) by dividing the blood pressure standard deviation by the mean BP then multiplying by 100, and 6) a variability score (from 0 [low] to 100 [high]) defined as the number of changes in BP of 5 mmHg or more, i.e., 100*(number of absolute BP change of each two successive measurements>5)/number of measurements.

Clinical characteristics were summarized using statistical descriptive statistics. Continuous variables were presented as median (95% confidence interval [CI] or interquartile range [IQR]) and categorical variables were presented count (%). The Mann-Whitney U test was used to compare continuous variables. The χ^2^ test with Yates’ correction was used for 2×2 contingency data, and Pearson’s χ^2^ test was used for contingency data for variables with more than two categories. Univariate Cox regression models were conducted based on cohorts of males and females, respectively, to identify the significant predictors of GAD. Significant univariate predictors of demographics, prior comorbidities, clinical and biochemical tests, medication prescriptions and BPV were used as input of a multivariate Cox analysis model, adjusted for demographics and comorbidities. Hazard ratios (HRs) with corresponding 95% CIs and P-values were reported. All significance tests were two-tailed and considered significant if P-values <0.001. Data analysis was performed using RStudio software (Version: 1.1.456) and Python (Version: 3.6).

## Results

### Baseline clinical characteristics and GAD incidence

This study included a total of 48023 (50% males) patients with a median follow-up of 224 (IQR: 217-229) months (**Supplementary Figure 1**). Among the 23964 male patients, 495 (incidence rate: 2.1%, median age: 70 (IQR: 57-79) years old) developed GAD. By contrast, females had a higher incidence rate, with 1687 of 24059 (incidence rate: 7.0%, median age: 68 (56-78) years old) developing GAD.

The clinical characteristics of the included patients are provided in **Table 1**. Compared with female patients, male patients were more likely to suffer from cardiovascular diseases (63.63% v.s. 52.97%, p<0.0001), kidney disease (29.89% v.s. 17.10%, p< 0.0001), and stroke (38.38% v.s. 28.30%, p<0.0001). They were more likely to be prescribed angiotensinogen-converting enzyme inhibitors (ACEI) (18.18% v.s. 11.20%) and other antihypertensive drugs (23.03% v.s. 4.82%) than female patients.

**Table 1.** Clinical characteristics of patients included in this cohort. * for p≤ 0.05, ** for p ≤ 0.01, *** for p ≤ 0.001

| **Characteristics** | **Males (N=23964; event: 495,**<br>**incidence rate: 2.07%;**<br>**mortality: 195, 39.4%)**<br>**Median (IQR);Max;N or Count(%)** | **Females (N=24059; event: 1687,**<br>**incidence rate: 7.01%;**<br>**mortality: 431, 25.68%)**<br>**Median (IQR);Max;N or Count(%)** | **P** |
| --- | --- | --- | --- |
| **Demographics** |  |  |  |
| Age of first BP test, years | 61.4(50.8-69.8);88.3;n=495 | 59.3 (49.3-69.3);89.0;n=1678 | 0.1453 |
| **Past comorbidity** |  |  |  |
| Cardiovascular | 347(70.10%) | 1209(72.05%) | 0.2253 |
| Respiratory | 315(63.63%) | 889(52.97%) | <0.0001*** |
| Kidney | 148(29.89%) | 287(17.10%) | <0.0001*** |
| Endocrine | 25(5.05%) | 83(4.94%) | 0.2588 |
| Diabetes mellitus | 79(15.95%) | 287(17.10%) | 0.1086 |
| Hypertension | 336(67.87%) | 1131(67.40%) | 0.3494 |
| Gastrointestinal | 275(55.55%) | 942(56.13%) | 0.2378 |
| Obesity | 3(0.60%) | 6(0.35%) | 0.8561 |
| Stroke | 190(38.38%) | 475(28.30%) | <0.0001*** |
| **Medications** |  |  |  |
| ACEI | 90(18.18%) | 188(11.20%) | 0.0025** |
| ARB | 2(0.40%) | 9(0.53%) | 0.8262 |
| Calcium channel blockers | 140(28.28%) | 379(22.58%) | 0.1477 |
| Beta blockers | 156(31.51%) | 537(32.00%) | 0.5235 |
| Diuretics for heart failure | 14(2.82%) | 51(3.03%) | 0.5597 |
| Diuretics for hypertension | 59(11.91%) | 197(11.74%) | 0.2936 |
| Nitrates | 74(14.94%) | 203(12.09%) | 0.8393 |
| Antihypertensive drugs | 114(23.03%) | 81(4.82%) | <0.0001*** |
| Anti-Diabetic drugs | 54(10.90%) | 175(10.42%) | 0.5855 |
| Statins and fibrates | 75(15.15%) | 258(15.37%) | 0.6512 |
| **Complete blood count** |  |  |  |
| Mean corpuscular volume, fL | 91.3(88.3-94.0);108.4;n=242 | 89.4(86.2-92.5);106.0;n=889 | 0.1994 |
| Basophil, x10<sup>9</sup>/L | 0.04(0.02-0.046);0.6;n=113 | 0.03(0.01-0.03);0.2;n=390 | 0.35 |
| Eosinophil, x10<sup>9</sup>/L | 0.1(0.1-0.2);1.7;n=132 | 0.1(0.1-0.2);8.8;n=485 | 0.3711 |
| Lymphocyte, x10<sup>9</sup>/L | 1.78(1.3-2.2);4.5;n=133 | 1.8(1.4-2.4);5.2;n=489 | 0.1135 |
| Monocyte, x10<sup>9</sup>/L | 0.5(0.4-0.7);1.5;n=132 | 0.4(0.3-0.6);1.5;n=486 | 0.6279 |
| Neutrophil, x10<sup>9</sup>/L | 4.3(3.5-6.55);14.3;n=132 | 4.1(3.2-5.53);20.4;n=485 | 0.6455 |
| White cell count, x10<sup>9</sup>/L | 7.16(6.0-8.95);24.0;n=243 | 6.8(5.6-8.3);23.8;n=892 | 0.787 |
| Mean corpuscular haemoglobin, pg | 31.1(29.85-32.1);38.5;n=242 | 30.5(29.1-31.5);35.6;n=889 | 0.8937 |
| Platelet, x10<sup>9</sup>/L | 229.0(191.0-265.0);419.0;n=243 | 244.0(209.0-293.0);563.0;n=891 | 0.431 |
| Reticulocyte, x10<sup>9</sup>/L | 31.9(28.8-60.9);90.0;n=3 | 60.9(44.0-91.77);162.0;n=17 | <0.0001*** |
| Red cell count, x10<sup>12</sup>/L | 4.65(4.34-5.01);6.68;n=241 | 4.31(4.04-4.6);6.5;n=889 | 0.3762 |
| Hematocrit, L/L | 0.43(0.4-0.45);0.54;n=226 | 0.38(0.4-0.4);0.5;n=836 | 0.9949 |
| **Renal and liver function tests** |  |  |  |
| Potassium, mmol/L | 4.15(3.89-4.5);6.44;n=337 | 4.2(3.9-4.47);8.9;n=1041 | 0.297 |
| Urate, mmol/L | 0.398(0.35-0.475);0.758;n=86 | 0.32(0.26-0.39);0.69;n=253 | 0.1253 |
| Albumin, g/L | 42.25(40.0-44.368);54.0;n=290 | 42.0(39.8-44.0);52.0;n=941 | 0.9327 |
| Sodium, mmol/L | 141.0(139.0-142.0);153.0;n=337 | 141.0(139.0-142.0);148.0;n=1041 | 0.5236 |
| Urea, mmol/L | 5.7(4.835-6.8);26.1;n=336 | 5.3(4.3-6.3);21.5;n=1039 | 0.9642 |
| Protein, g/L | 74.0(70.69-77.35);89.0;n=287 | 74.0(71.0-78.0);90.2;n=939 | 0.6126 |
| Creatinine, umol/L | 98.0(88.0-111.0);311.0;n=337 | 75.0(67.0-85.0);305.0;n=1041 | <0.0001*** |
| Alkaline phosphatase, U/L | 75.0(60.84-92.0);218.0;n=248 | 77.0(60.0-93.5);294.0;n=815 | 0.7759 |
| Aspartate transaminase, U/L | 23.0(17.0-27.0);416.0;n=68 | 21.0(18.0-26.0);770.0;n=214 | 0.544 |
| Alanine transaminase, U/L | 23.0(17.0-35.0);275.0;n=225 | 19.0(14.0-27.0);570.9;n=740 | 0.0023** |
| Bilirubin, umol/L | 10.0(7.5-13.0);41.0;n=250 | 9.0(6.3874-11.3);45.2;n=824 | 0.3542 |
| **Glycemic and lipid profile** |  |  |  |
| Triglyceride, mmol/mol | 1.6(1.1-2.3);6.3;n=156 | 1.4(0.97-2.0);13.97;n=546 | 0.6491 |
| LDL, mmol/mol | 3.3(2.8-3.7);5.8;n=116 | 3.2(2.6-3.9);6.5409;n=373 | 0.3887 |
| HDL, mmol/mol | 1.2(1.02-1.4);2.1;n=118 | 1.4(1.195-1.66);2.78;n=390 | 0.3572 |
| HbA1c, g/dL | 13.9(12.9-15.1);17.8;n=215 | 12.8(11.7-13.6);16.7;n=788 | 0.0031** |
| Cholesterol, mmol/L | 5.2(4.6-5.7);7.95;n=156 | 5.4(4.8-6.1);8.95;n=548 | 0.128 |
| Fast glucose, mmol/L | 5.8(5.2-7.5);24.7;n=240 | 5.6(5.1-7.0);28.2;n=787 | 0.9374 |
| **Diastolic blood pressure measures** |  |  |  |
| Number of measurements | 7(5-10);25;n=495 | 7(6-10);23;n=1678 | 0.1225 |
| Baseline, mm Hg | 77.0(68.0-84.5);122.0;n=495 | 72.0(65.0-80.0);110.0;n=1678 | 0.041* |
| Latest, mm Hg | 74.0(66.0-81.0);135.0;n=495 | 70.0(63.0-78.0);128.0;n=1678 | 0.6643 |
| Maximum, mm Hg | 89.0(81.5-97.0);135.0;n=495 | 86.0(79.0-93.0);128.0;n=1678 | 0.8432 |
| Minimal, mm Hg | 63.0(56.0-70.0);100.0;n=495 | 58.0(52.0-66.0);98.0;n=1678 | 0.0323* |
| Mean, mm Hg | 75.4(69.8-81.6);106.4286;n=495 | 71.9(66.2-77.2);102.0;n=1678 | 0.0065** |
| Median, mm Hg | 75.5(69.5-81.0);110.0;n=495 | 71.5(66.0-77.0);102.0;n=1678 | 0.0234* |
| Variance | 54.3(31.4-88.9);512.0;n=495 | 56.1(33.4-84.3);924.5;n=1678 | 0.2326 |
| SD | 7.4(5.6-9.4);22.6;n=495 | 7.5(5.8-9.2);30.4;n=1678 | 0.5344 |
| RMS | 75.9(70.3-81.9);106.6;n=495 | 72.3(66.6-77.6);102.1;n=1678 | 0.0422* |
| CV | 0.09(0.07-0.12);0.25;n=495 | 0.1001(0.073-0.1);0.31;n=1678 | <0.0001*** |
| Variability score | 57.1(46.2-66.7);93.8;n=495 | 55.8(47.4-66.2);92.3;n=1678 | 0.5416 |
| **Systolic blood pressure measures** |  |  |  |
| Number of measurements |  |  |  |
| Baseline, mm Hg | 135.0(120.0-150.0);217.0;n=495 | 132.0(117.0-147.0);225.0;n=1678 | 0.1594 |
| Latest, mm Hg | 132.0(121.0-143.0);209.0;n=495 | 131.0(119.0-142.0);234.0;n=1678 | 0.7247 |
| Maximum, mm Hg | 156.0(144.0-169.0);233.0;n=495 | 157.0(140.0-173.0);246.0;n=1678 | 0.7289 |
| Minimal, mm Hg | 111.0(102.0-121.0);164.0;n=495 | 109.0(101.0-120.0);183.0;n=1678 | 0.1322 |
| Mean, mm Hg | 134.0(125.4-142.4);191.6;n=495 | 132.5(123.3-141.0);193.3;n=1678 | 0.3252 |
| Median, mm Hg | 133.0(125.0-142.0);187.0;n=495 | 132.0(122.5-141.0);195.0;n=1678 | 0.2253 |
| Variance | 165.7(96.9-255.0);1512.5;n=495 | 161.8(91.7-256.7);2964.5;n=1678 | 0.1806 |
| SD | 12.9(9.8-15.9);38.9;n=495 | 12.7(9.6-16.0);54.4;n=1678 | 0.2178 |
| RMS | 134.7(126.2-143.0);192.4;n=495 | 133.4(124.0-141.9);193.6;n=1678 | 0.2509 |
| CV | 0.09(0.07-0.11);0.2;n=495 | 0.09(0.07-0.12);0.27;n=1678 | 0.2264 |
| Variability score | 70.0(60.0-77.0);94.4;n=495 | 70.6(57.1-78.6);93.8;n=1678 | 0.7872 |

ACEI: angiotensinogen-converting enzyme inhibitor; ARB: angiotensin receptor blocker; LDL: low density lipoprotein cholesterol; HDL: high density lipoprotein cholesterol; SD: standard deviation; RMS: root mean square; CV: coefficient of variation

Nevertheless, female patients had higher reticulocyte level (median: 60.9, IQR: 44.0-91.77 v.s. median: 31.9, IQR: 28.8-60.9, p<0.0001), lower creatinine level (median: 75.0, IQR: 67.0-85.0 v.s. median: 98.0, IQR: 88.0-111.0, p<0.0001), lower alanine transaminase amount (median: 19.0, IQR: 14.0-27.0 v.s. median: 23.0, IQR: 17.0-35.0, p=0.0023), lower HbA1c level (median: 12.8, IQR: 11.7-13.6 v.s. median: 13.9, IQR: 12.9-15.1, p=0.0031). As regards to diastolic BP measurements, female patients had a lower mean (median: 58.0, IQR: 52.0-66.0 v.s. median: 63.0, IQR: 56.0-70.0, p=0.0323), median (median: 71.9, IQR: 66.2-77.2 v.s. median: 75.4, IQR: 69.8-81.6, p=0.0065), RMS (median: 72.3, IQR: 66.6-77.6 v.s. median: 75.9, IQR: 70.3-81.9, p=0.0422), and CV (median: 0.1001, IQR: 0.073-0.1 v.s. median: 0.09, IQR: 0.07-0.12, p<0.0001).

### Incidence of GAD on follow-up and significant predictors

The age-specific incidences of GAD among male and female subgroups are shown in **Figure 1**. The number of female patients developing GAD was more than double that of male patients among those over 30 years of age. Kaplan-Meier survival curves in **Figure 2** show that females had a higher risk of developing GAD than males.

**Figure 1.**
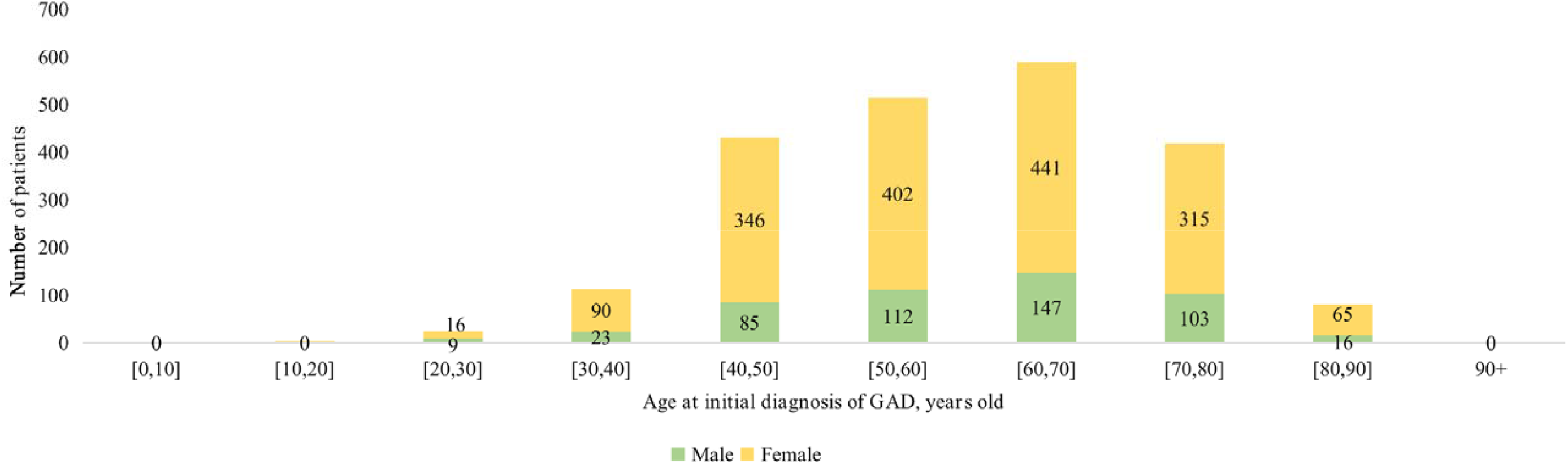
Age-specific incidence of GAD among male and female subgroups.

**Figure 2.**
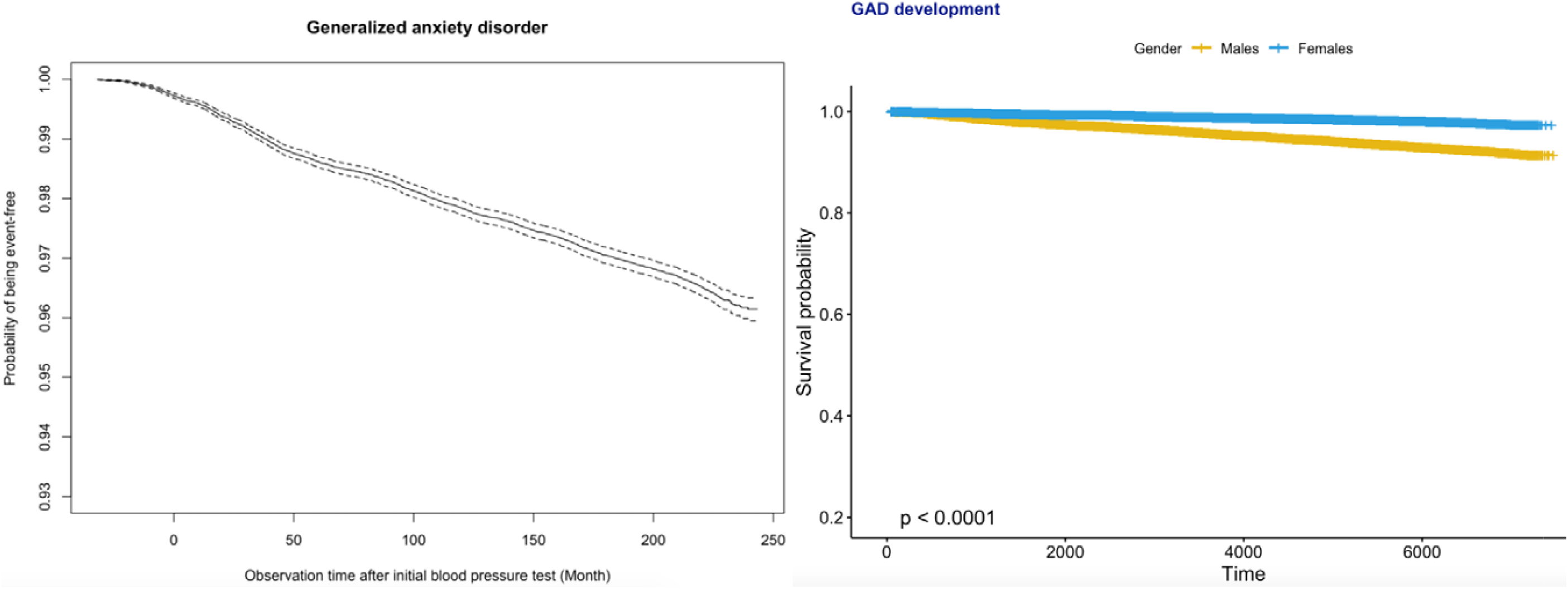
Kaplan-Meier survival curves of incident GAD development for the whole cohort (top) and stratified by gender (bottom).

Univariate Cox regression demonstrated the following significant predictors for incident GAD:

1. Demographics, namely, female gender (HR: 0.30, 95% CI: [0.27,0.33], p<0.0001***) and older age (HR: 1.23, 95% CI: [1.19,2.03], p<0.0001). The specific risks differed between age groups: [40,50] years old (HR: 1.42, 95% CI: [1.27,1.57], p<0.0001), [50,60] years old (HR: 1.30, 95% CI: [1.18,1.43], p<0.0001), [60,70] years old (HR: 1.11, 95% CI: [1.01,1.22], p=0.0008), [70,80] years old (HR: 1.71, 95% CI: [1.64,1.79], p<0.0001), [80,90] years old (HR: 1.46, 95% CI: [1.37,1.57], p<0.0001);
2. Past history of cardiovascular diseases (HR: 4.00, 95% CI: [3.64,4.39], p<0.0001), respiratory diseases (HR: 1.33, 95% CI: [1.22,1.45], p<0.0001), diabetes mellitus (HR: 1.17, 95% CI: [1.05,1.31], p=0.0062), hypertension (HR: 1.17, 95% CI: [1.07,1.28], p=0.0008), and gastrointestinal disorders (HR: 1.90, 95% CI: [1.74,2.07], p<0.0001);
3. Laboratory parameters, namely, lower neutrophil (HR: 0.35, 95% CI: [0.24,0.50], p<0.0001), less white cell count (HR: 0.92, 95% CI: [0.89,0.95], p<0.0001), lower mean corpuscular haemoglobin level (HR: 0.94, 95% CI: [0.92,0.96], p<0.0001), higher red cell count (HR: 1.001[1.001,1.002], p=0.0002), lower urate level (HR: 0.03, 95% CI: [0.01,0.07], p<0.0001), higher albumin level (HR: 1.04, 95% CI: [1.03,1.06], p<0.0001), lower urea level (HR: 0.87, 95% CI: [0.85,0.90], p<0.0001), lower creatinine level (HR: 0.98, 95% CI: [0.979,0.982], p<0.0001), lower alkaline phosphatase level (HR: 0.995, 95% CI: [0.993,0.997], p<0.0001), lower bilirubin level (HR: 0.97, 95% CI: [0.95,0.98], p<0.0001), higher high-density lipoprotein (HDL) level (HR: 1.54, 95% CI: [1.25,1.91], p=0.0001), and lower fasting glucose level (HR: 0.96, 95% CI: [0.93,0.98], p: 0.0007);
4. Diastolic blood pressure measures, namely, higher baseline value (HR: 1.49, 95% CI: [1.08,2.45], p<0.0001), higher maximum value (HR: 1.19, 95% CI: [1.06,1.54], p<0.0001), higher minimum value (HR: 1.23, 95% CI: [1.08,2.06], p<0.0001), larger SD (HR: 1.18, 95% CI: [1.03,1.95], p=0.0008), larger CV (HR: 1.13, 95% CI: [1.05,1.38], p=0.0002), and larger variability score (HR: 1.16, 95% CI: [1.04,1.85], p=0.0003);
5. Larger values of all systolic blood pressure measures (baseline, latest, maximum, minimum, mean, median, variance, SD, RMS, CV, variability score) (HR>1, p <0.001).

In addition, the significant univariate predictors of all-cause mortality after GAD presentation were also identified (**Table 2**). Significant univariate predictors (p<0.05) were entered into a multivariate Cox regression model, with most of the above univariate predictors remaining significant (**Table 3**). Next, we further analyzed different BP values in patients who developed GAD with age stratification (**Figures 3 and 4**; **Supplementary Table 2**). There is an age-related increase in mean, median and measures of variability for both diastolic and systolic BP.

**Table 2.** Univariate Cox analysis to predict incident GAD and mortality. * for p≤ 0.05, ** for p ≤ 0.01, *** for p ≤ 0.001

| Characteristics | GAD (N=2173)<br>HR [95% CI] |  | Mortality (N=626)<br>HR [95% CI] | P |
| --- | --- | --- | --- | --- |
| **Demographics** |  |  |  |  |
| Male gender | 0.30[0.27,0.33] | <0.0001*** | 1.74[1.69,1.79] | <0.0001*** |
| Age at first blood pressure test, years | 1.23[1.19,2.03] | <0.0001*** | 1.10[1.10,1.11] | <0.0001*** |
| [0,10] | - | - | - | - |
| [10,20] | 0.31[0.10,0.97] | 0.0446 * | 0.01[0.00,0.07] | <0.0001*** |
| [20,30] | 0.54[0.36,0.79] | 0.0019 ** | 0.13[0.10,0.17] | <0.0001*** |
| [30,40] | 1.10[0.91,1.33] | 0.3140 | 0.06[0.05,0.08] | <0.0001*** |
| [40,50] | 1.42[1.27,1.57] | <0.0001*** | 0.11[0.10,0.12] | <0.0001*** |
| [50,60] | 1.30[1.18,1.43] | <0.0001*** | 0.25[0.24,0.27] | <0.0001*** |
| [60,70] | 1.11[1.01,1.22] | 0.0008** | 0.97[0.94,1.00] | 0.0749 |
| [70,80] | 1.71[1.64,1.79] | <0.0001*** | 3.51[3.41,3.61] | <0.0001*** |
| [80,90] | 1.46[1.37,1.57] | <0.0001*** | 6.00[5.78,6.23] | <0.0001*** |
| 90+ | - | 0.9640 | 8.76[7.75,9.89] | <0.0001*** |
| **Past comorbidity** |  |  |  |  |
| Cardiovascular | 4.00[3.64,4.39] | <0.0001*** | 2.18[2.12,2.24] | <0.0001*** |
| Respiratory | 1.33[1.22,1.45] | <0.0001*** | 3.90[3.77,4.02] | <0.0001*** |
| Kidney | 0.87[0.78,0.96] | 0.0076 ** | 2.15[2.09,2.22] | <0.0001*** |
| Endocrine | 1.12[0.92,1.36] | 0.2500 | 2.08[1.97,2.20] | <0.0001*** |
| Diabetes mellitus | 1.17[1.05,1.31] | 0.0062 ** | 0.96[0.92,1.00] | 0.0385 * |
| Hypertension | 1.17[1.07,1.28] | 0.0008 *** | 0.87[0.84,0.90] | <0.0001*** |
| Gastrointestinal | 1.90[1.74,2.07] | <0.0001*** | 1.20[1.17,1.23] | <0.0001*** |
| Obesity | 2.07[1.08,3.98] | 0.0296 * | 0.39[0.25,0.62] | <0.0001*** |
| Stroke | 1.07[0.98,1.17] | 0.1410 | 1.94[1.89,2.00] | <0.0001*** |
| **Medications** |  |  |  |  |
| ACEI | 0.66[0.59,0.75] | <0.0001*** | 1.80[1.74,1.86] | <0.0001*** |
| ARB | 0.80[0.44,1.45] | 0.4600 | 1.50[1.29,1.75] | <0.0001*** |
| Calcium channel blockers | 0.70[0.64,0.78] | <0.0001*** | 1.85[1.80,1.91] | <0.0001*** |
| Beta blockers | 1.20[1.10,1.32] | 0.0001 *** | 1.06[1.03,1.10] | 0.0001 *** |
| Diuretics for heart failure | 0.67[0.52,0.85] | 0.0013 ** | 3.98[3.79,4.18] | <0.0001*** |
| Diuretics for hypertension | 0.84[0.73,0.95] | 0.0073 ** | 1.40[1.35,1.45] | <0.0001*** |
| Nitrates | 1.05[0.93,1.20] | 0.4130 | 1.91[1.84,1.98] | <0.0001*** |
| Antihypertensive drugs | 0.72[0.62,0.84] | <0.0001*** | 2.23[2.15,2.31] | <0.0001*** |
| Antidiabetic drugs | 0.59[0.51,0.67] | <0.0001*** | 1.40[1.35,1.45] | <0.0001*** |
| Statins and fibrates | 1.11[0.98,1.24] | 0.0908 . | 1.06[1.02,1.10] | 0.0068 ** |
| **Complete blood count** |  |  |  |  |
| Mean corpuscular volume, fL | 0.995[0.988,1.002] | 0.1930 | 1.021[1.018,1.024] | <0.0001*** |
| Basophil, x10<sup>9</sup>/L | 0.20[0.02,1.68] | 0.1370 | 2.00[1.05,3.79] | 0.0339 * |
| Eosinophil, x10<sup>9</sup>/L | 0.81[0.54,1.22] | 0.3160 | 1.11[1.02,1.21] | 0.0178 * |
| Lymphocyte, x10<sup>9</sup>/L | 1.03[1.01,1.06] | 0.0161 * | 0.66[0.63,0.68] | <0.0001*** |
| Monocyte, x10<sup>9</sup>/L | 1.88[0.33,10.83] | 0.4800 | 1.04[0.65,1.66] | 0.871 |
| Neutrophil, x10<sup>9</sup>/L | 0.35[0.24,0.50] | <0.0001*** | 2.10[1.96,2.26] | <0.0001*** |
| White cell count, x10<sup>9</sup>/L | 0.92[0.89,0.95] | <0.0001*** | 1.08[1.08,1.09] | <0.0001*** |
| Mean corpuscular haemoglobin, pg | 0.94[0.92,0.96] | <0.0001*** | 1.06[1.06,1.06] | <0.0001*** |
| Platelet, x10<sup>9</sup>/L | 1.00[0.98,1.01] | 0.6170 | 1.03[1.03,1.04] | <0.0001*** |
| Reticulocyte, x10<sup>9</sup>/L | 11.70[0.38,357.50] | 0.1590 | 1.78[0.89,3.56] | 0.105 |
| Red cell count, x10<sup>12</sup>/L | 1.001[1.001,1.002] | 0.0002 *** | 0.998[0.998,0.998] | <0.0001*** |
| Hematocrit, L/L | 1.003[0.993,1.013] | 0.5270 | 0.999[0.996,1.001] | 0.312 |
| Basophil, x10<sup>9</sup>/L | 0.97[0.88,1.06] | 0.4620 | 0.57[0.55,0.59] | <0.0001*** |
| Eosinophil, x10<sup>9</sup>/L | 0.63[0.19,2.07] | 0.4490 | 0.50[0.12,0.81] | <0.0001*** |
| **Renal and liver function test** |  |  |  |  |
| Potassium, mmol/L | 0.88[0.79,0.97] | 0.0102 * | 0.95[0.92,0.98] | 0.0026 ** |
| Urate, mmol/L | 0.03[0.01,0.07] | <0.0001*** | 10.65[8.23,13.77] | <0.0001*** |
| Albumin, g/L | 1.04[1.03,1.06] | <0.0001*** | 0.88[0.87,0.88] | <0.0001*** |
| Sodium, mmol/L | 1.02[1.00,1.04] | 0.0196 * | 0.948[0.943,0.952] | <0.0001*** |
| Urea, mmol/L | 0.87[0.85,0.90] | <0.0001*** | 1.111[1.108,1.114] | <0.0001*** |
| Protein, g/L | 1.01[1.00,1.02] | 0.0528. | 0.95[0.95,0.95] | <0.0001*** |
| Creatinine, umol/L | 0.98[0.979,0.982] | <0.0001*** | 1.003[1.003,1.003] | <0.0001*** |
| Alkaline phosphatase, U/L | 0.995[0.993,0.997] | <0.0001*** | 1.002[1.002,1.002] | <0.0001*** |
| Aspartate transaminase, U/L | 1.000[0.998,1.001] | 0.6580 | 1.001[1.000,1.001] | <0.0001*** |
| Alanine transaminase, U/L | 0.999[0.997,1.001] | 0.4250 | 0.998[0.997,0.999] | <0.0001*** |
| Bilirubin, umol/L | 0.97[0.95,0.98] | <0.0001*** | 1.007[1.005,1.009] | <0.0001*** |
| **Glycemic and lipid profile** |  |  |  |  |
| Triglyceride, mmol/mol | 0.98[0.92,1.04] | 0.4300 | 0.97[0.95,0.99] | 0.0024 ** |
| LDL, mmol/mol | 1.03[0.93,1.13] | 0.6050 | 0.91[0.88,0.95] | <0.0001*** |
| HDL, mmol/mol | 1.54[1.25,1.91] | 0.0001 *** | 0.82[0.75,0.90] | <0.0001*** |
| HbA1c, g/dL | 1.01[1.00,1.03] | 0.0613 | 0.98[0.98,0.98] | <0.0001*** |
| Cholesterol, mmol/L | 1.04[0.98,1.11] | 0.2230 | 0.92[0.90,0.94] | <0.0001*** |
| Fast glucose, mmol/L | 0.96[0.93,0.98] | 0.0007 *** | 1.05[1.05,1.06] | <0.0001*** |
| **Diastolic blood pressure measures** |  |  |  |  |
| Number of measurements | 0.89[0.18,1.23] | 0.1352 | 0.53[0.42,1.71] | 0.2342 |
| Baseline, mm Hg | 1.49[1.08,2.45] | <0.0001*** | 1.03[1.01,1.24] | <0.0001*** |
| Latest, mm Hg | 1.001[0.997,1.004] | 0.6870 | 1.07[1.02,1.28] | <0.0001*** |
| Maximum, mm Hg | 1.19[1.06,1.54] | <0.0001*** | 1.04[1.02,1.08] | <0.0001*** |
| Minimal, mm Hg | 1.23[1.08,2.06] | <0.0001*** | 1.09[1.01,1.2] | 0.0442 * |
| Mean, mm Hg | 1.03[1.01,1.39] | 0.0011 ** | 1.1[1.03,1.7] | <0.0001*** |
| Median, mm Hg | 1.04[1.01,1.53] | 0.0026 ** | 1.28[1.16,1.98] | <0.0001*** |
| Variance | 1.002[1.001,1.003] | 0.0491 * | 1.003[1.003,1.003] | <0.0001*** |
| SD | 1.18[1.03,1.95] | 0.0008 *** | 1.06[1.05,1.06] | <0.0001 *** |
| RMS | 1.23[1.09,1.6] | 0.0078 ** | 1.18[1.02,1.93] | <0.0001 *** |
| CV | 1.13[1.05,1.38] | 0.0002 *** | 40.89[28.15,59.38] | <0.0001 *** |
| Variability score | 1.16[1.04,1.85] | 0.0003 *** | 1.001[1.000,1.002] | 0.0082 ** |
| **Systolic blood pressure measures** |  |  |  |  |
| Number of measurements |  |  |  |  |
| Baseline, mm Hg | 1.25[1.04,1.93] | <0.0001 *** | 1.015[1.014,1.015] | <0.0001 *** |
| Latest, mm Hg | 1.34[1.12,1.88] | <0.0001 *** | 1.009[1.008,1.010] | <0.0001 *** |
| Maximum, mm Hg | 1.15[1.04,1.33] | <0.0001 *** | 1.009[1.009,1.010] | <0.0001 *** |
| Minimal, mm Hg | 1.04[1.01,1.07] | <0.0001 *** | 1.022[1.021,1.023] | <0.0001 *** |
| Mean, mm Hg | 1.26[1.03,1.56] | <0.0001 *** | 1.031[1.030,1.032] | <0.0001 *** |
| Median, mm Hg | 1.16[1.04,1.29] | <0.0001 *** | 1.030[1.029,1.031] | <0.0001 *** |
| Variance | 1.04[1.01,1.1] | <0.0001 *** | 1.001[1.001,1.001] | <0.0001 *** |
| SD | 1.17[1.07,1.85] | <0.0001 *** | 1.061[1.059,1.063] | <0.0001 *** |
| RMS | 1.59[1.18,1.99] | <0.0001 *** | 1.031[1.030,1.032] | <0.0001 *** |
| CV | 1.07[1.02,1.23] | <0.0001 *** | 629.90[424.60,934.40] | <0.0001 *** |
| Variability score | 1.26[1.04,1.82] | 0.0008 *** | 1.000[0.999,1.001] | 0.8170 |

ACEI: angiotensinogen-converting enzyme inhibitor; ARB: angiotensin receptor blocker; LDL: low density lipoprotein cholesterol; HDL: high density lipoprotein cholesterol; SD: standard deviation; RMS: root mean square; CV: coefficient of variation

**Table 3.** Multivariate Cox regression analysis to predict incident GAD. * for p≤ 0.05, ** for p ≤ 0.01, *** for p ≤ 0.001

|  | HR [95% CI] | Z value | P |
| --- | --- | --- | --- |
| **Demographics** |  |  |  |
| Male gender | 0.23[0.11,0.48] | -3.93 | 0.0001*** |
| Age at first blood pressure test, years | 1.04[1.02,1.07] | 0.15 | 0.0029** |
| [20,30] | 1.24[1.26,5.98] | 0.27 | 0.0019** |
| [30,40] | 1.07[1.02,2.14] | 0.56 | 0.0034** |
| [40,50] | 1.98[1.38,10.24] | 0.82 | <0.0001*** |
| [50,60] | 1.47[1.22,9.92] | 0.39 | 0.0056** |
| [60,70] | 1.48[1.05,4.88] | 0.63 | <0.0001*** |
| [70,80] | 1.06[1.01,16.38] | 0.004 | 0.0069** |
| **Past comorbidities** |  |  |  |
| Cardiovascular | 3.69[1.99,6.84] | 4.16 | <0.0001*** |
| Respiratory | 1.68[0.96,2.94] | 1.83 | 0.0670. |
| Kidney | 1.72[0.98,3.04] | 1.88 | 0.0603. |
| Diabetes mellitus | 2.04[1.14,3.68] | 2.39 | 0.0169* |
| Hypertension | 1.42[0.69,2.88] | 0.96 | 0.3390 |
| Gastrointestinal | 1.24[0.74,2.07] | 0.82 | 0.4101 |
| **Medications** |  |  |  |
| ACEI | 0.95[0.54,1.68] | -0.18 | 0.8545 |
| Calcium channel blockers | 0.62[0.36,1.09] | -1.67 | 0.0947. |
| Beta blockers | 1.54[1.12,2.59] | 1.65 | 0.0016** |
| Diuretics for heart failure | 1.68[0.77,3.67] | 1.31 | 0.1915 |
| Diuretics for hypertension | 0.92[0.51,1.67] | -0.27 | 0.7879 |
| Antihypertensive drugs | 0.77[0.36,1.68] | -0.66 | 0.5125 |
| Antidiabetic drugs | 0.39[0.20,1.79] | -2.61 | 0.242 |
| **Complete blood count** |  |  |  |
| Neutrophil, x10<sup>9</sup>/L | 0.98[0.85,1.14] | -0.21 | 0.8336 |
| White cell count, x10<sup>9</sup>/L | 1.00[0.87,1.14] | -0.03 | 0.9728 |
| Mean corpuscular haemoglobin, pg | 1.07[0.95,1.21] | 1.09 | 0.2749 |
| Red blood count, x10<sup>12</sup>/L | 1.08[0.58,1.99] | 0.23 | 0.8179 |
| **Renal and liver function test** |  |  |  |
| Urate, mmol/L | 0.21[0.02,2.51] | -1.24 | 0.2161 |
| Albumin, g/L | 1.00[0.93,1.07] | -0.14 | 0.8891 |
| Urea, mmol/L | 0.97[0.83,1.13] | -0.42 | 0.6770 |
| Creatinine, umol/L | 1.00[0.98,1.01] | -0.65 | 0.5184 |
| Alkaline phosphatase, U/L | 0.99[0.98,1.00] | -1.44 | 0.1498 |
| Bilirubin, umol/L | 1.02[1.00,1.04] | 2.22 | 0.0264* |
| **Glycemic and lipid profile** |  |  |  |
| HDL, mmol/mol | 1.00[0.50,1.98] | -0.01 | 0.9921 |
| Fasting glucose, mmol/L | 1.04[0.98,1.11] | 1.32 | 0.1885 |
| **Diastolic blood pressure measures** |  |  |  |
| Baseline, mm Hg | 1.03[1.01,1.07] | 1.69 | <0.0001*** |
| Maximum, mm Hg | 0.98[0.91,1.06] | -0.48 | 0.6316 |
| Minimal, mm Hg | 1.04[1.01,1.14] | 0.90 | 0.0071** |
| Mean, mm Hg | 1.10[1.02,1.46] | 0.61 | 0.0011** |
| Median, mm Hg | 0.91[0.74,1.12] | -0.9 | 0.3679 |
| SD | 0.70[0.40,1.24] | -1.23 | 0.2204 |
| CV | 1.05[1.01,1.12] | 1.05 | 0.0659* |
| Variability score | 1.02[1.00,1.04] | 2.01 | 0.0047** |
| **Systolic blood pressure measures** |  |  |  |
| Baseline, mm Hg | 1.15[1.08,1.33] | 0.40 | <0.0001*** |
| Latest, mm Hg | 1.01[0.99,1.03] | 1.18 | 0.2389 |
| Maximum, mm Hg | 1.00[0.96,1.04] | 0.09 | 0.9302 |
| Minimal, mm Hg | 1.04[1.01,1.19] | 2.27 | 0.003** |
| Mean, mm Hg | 0.49[0.02,11.33] | -0.44 | 0.6579 |
| Median, mm Hg | 1.04[0.93,1.16] | 0.68 | 0.4949 |
| Variance | 1.00[0.99,1.01] | 0.38 | 0.7017 |
| SD | 1.08[0.70,1.67] | 0.36 | 0.7162 |
| RMS | 1.96[0.09,44.35] | 0.42 | 0.6719 |
| CV | 1.04[1.01,1.09] | 1.14 | 0.2540 |
| Variability score | 1.03[1.01,1.05] | 1.27 | 0.7872 |

ACEI: angiotensinogen-converting enzyme inhibitor; HDL: high density lipoprotein cholesterol; SD: standard deviation; RMS: root mean square; CV: coefficient of variation

**Figure 3.**
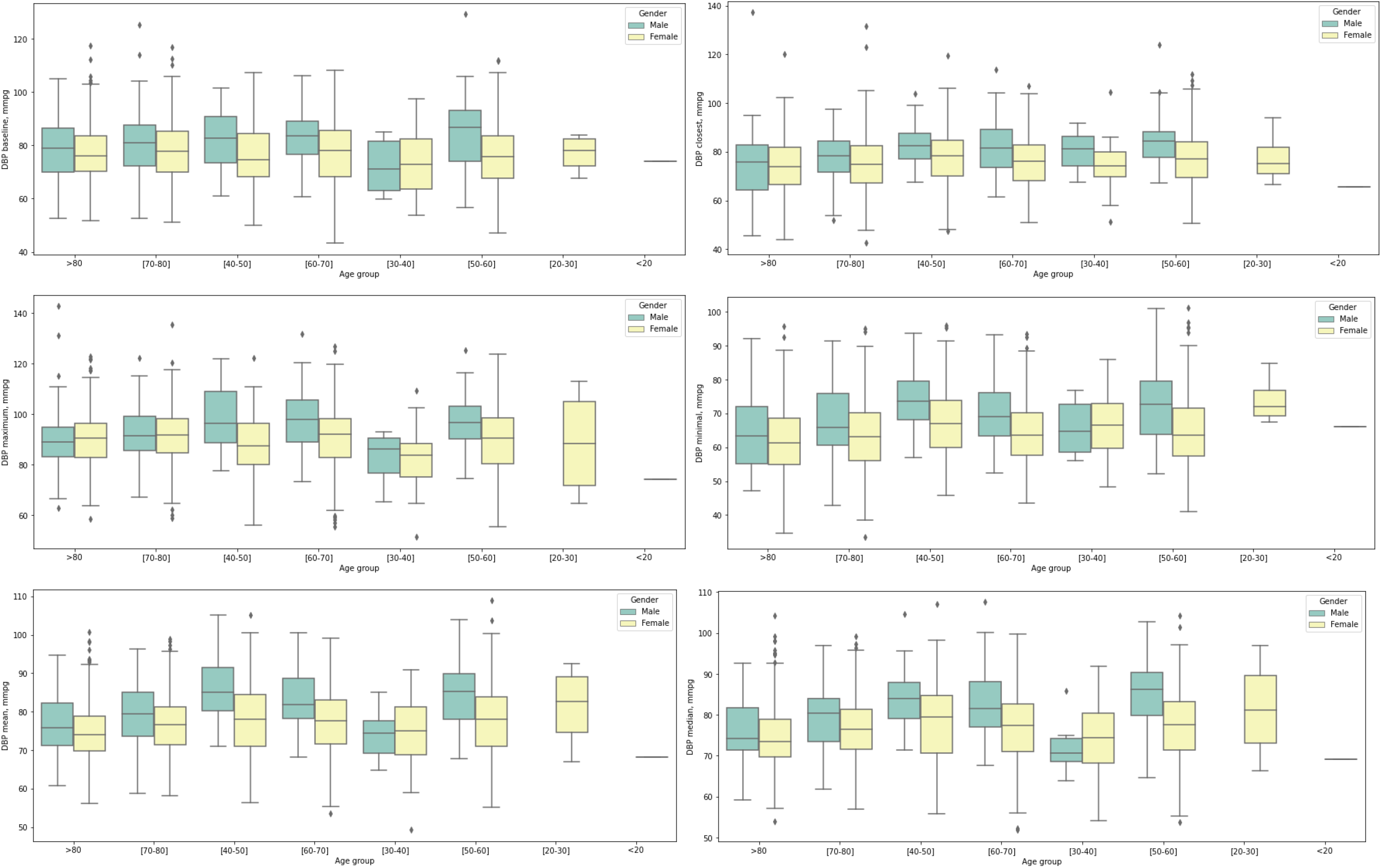
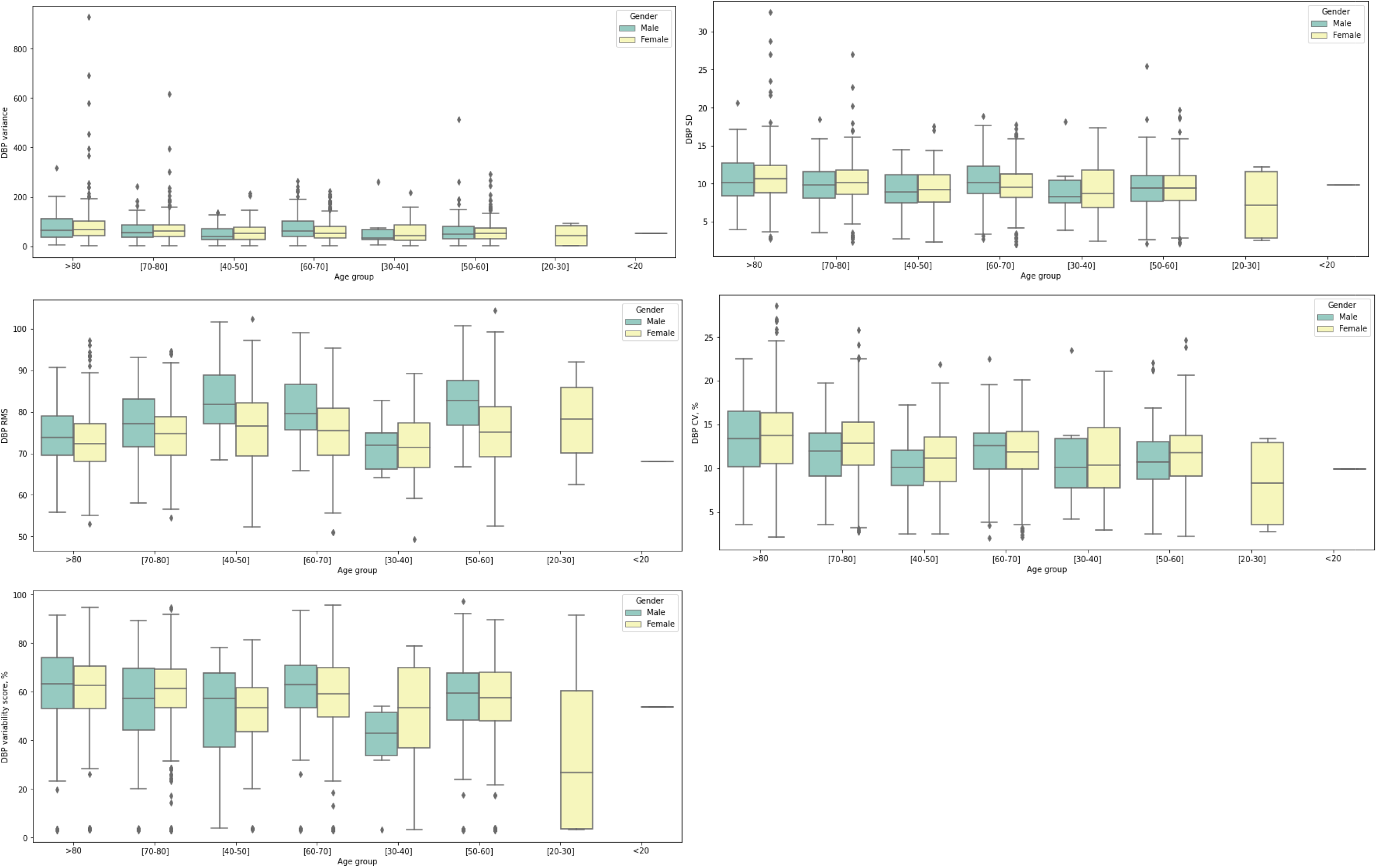
Box plots showing diastolic blood pressure (DBP) measures stratified by gender and age at GAD presentation.

**Figure 4.**
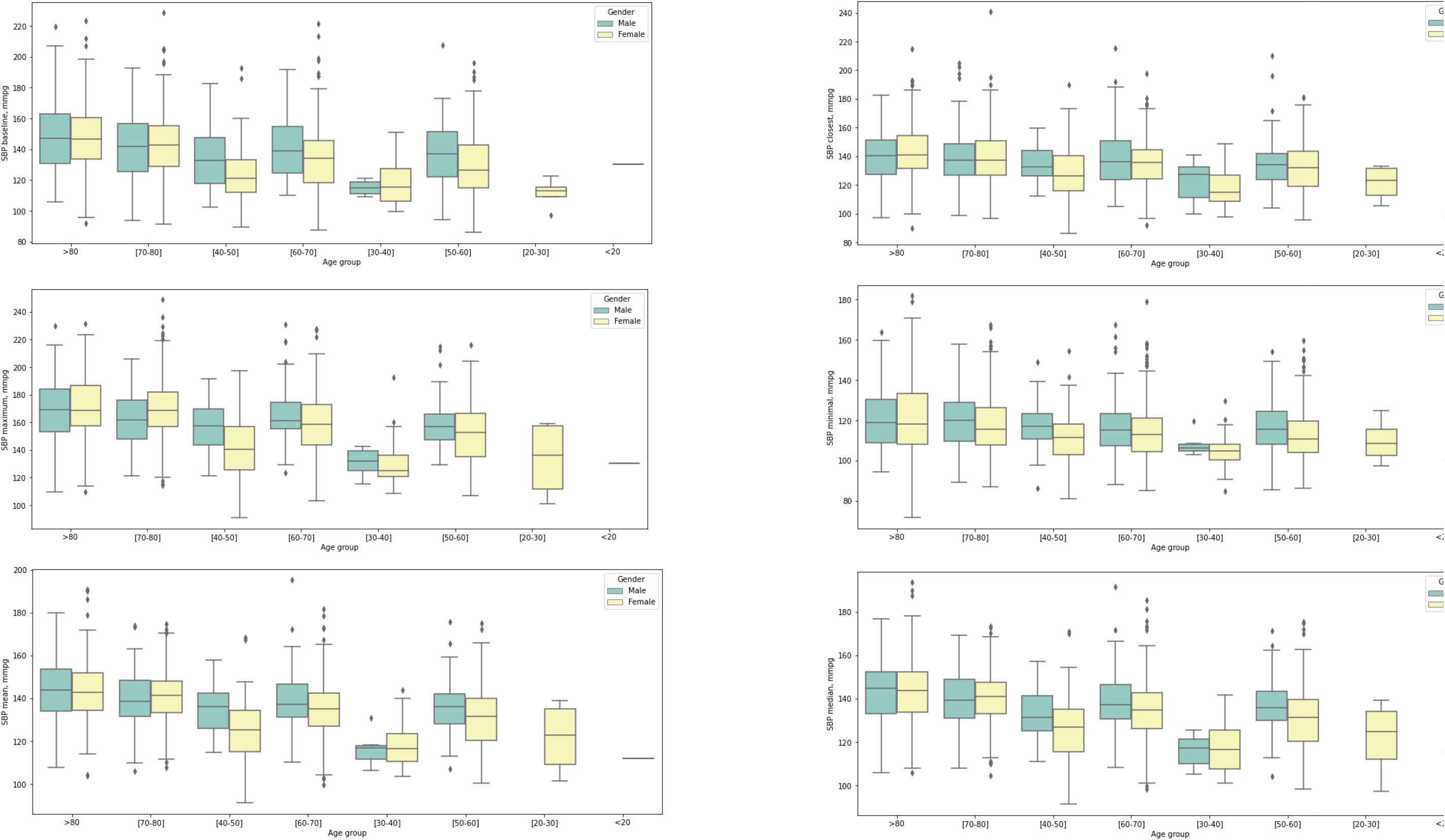
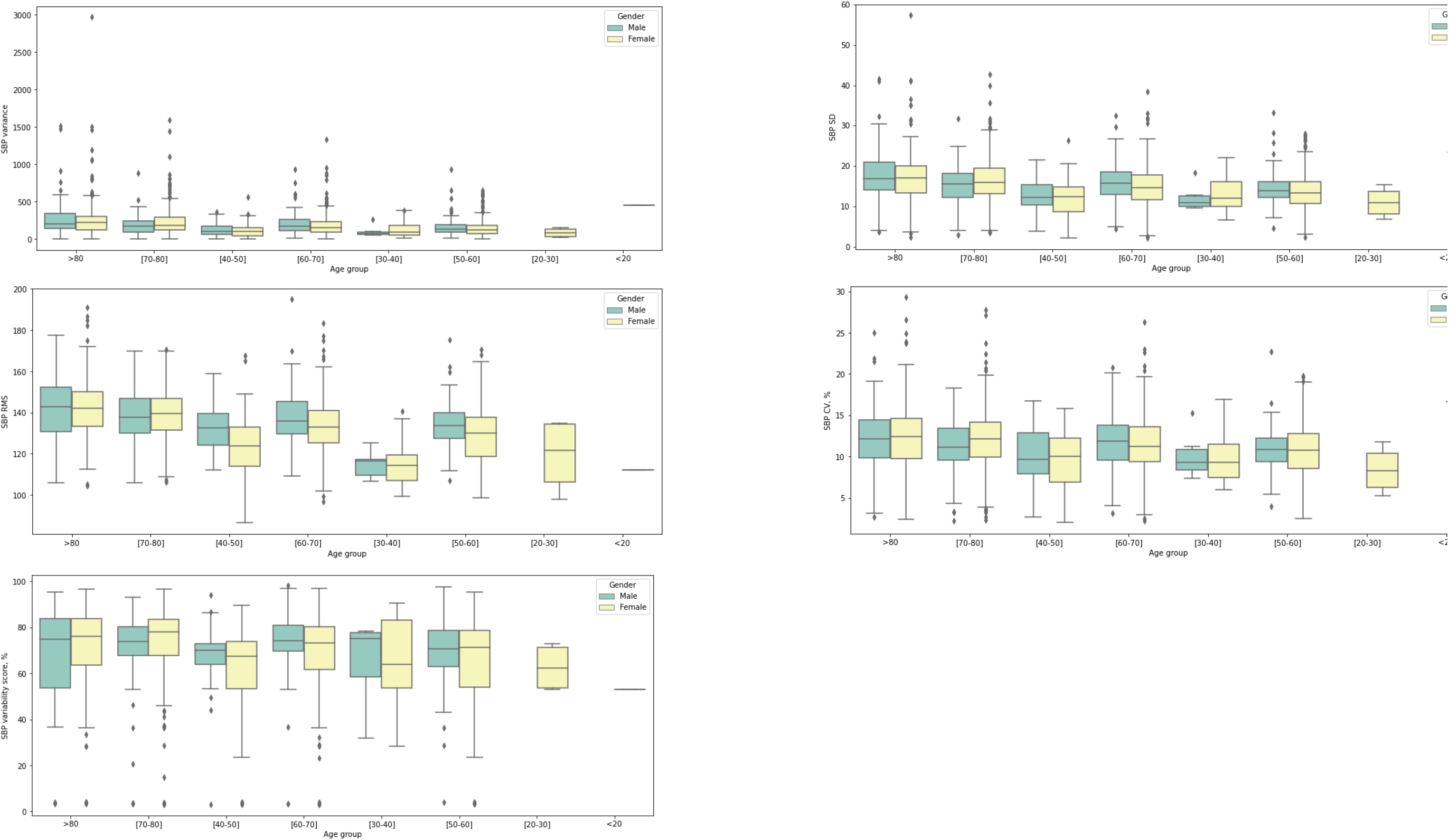
Box plots for showing systolic blood pressure (SBP) measures stratified by gender and age at GAD presentation.

### Comparisons of BP measurements before and after GAD development

Gender-specific blood pressure measure changes before and after initial GAD presentation were identified as shown in **Table 4**. Regarding systolic BP, males had larger variance (median: 172.4, IQR: 99.5-262.7 v.s. median: 165.7, IQR: 96.9-255.0, p<0.0001), larger RMS (median: 137.1, IQR: 125.5-144.0 v.s. median: 134.7, IQR: 126.2-143.0, p=0.0489), and larger variability score (median: 73.6, IQR: 60.5-80.3 v.s. median: 70.0, IQR: 60.0-77.0, p=0.0332) after initial presentation of GAD diseases; and females had larger variance (median: 174.2, IQR: 88.8-259.2 v.s. median 161.8, IQR: 91.7-256.7, p<0.0001) and larger variability score (median: 74.4, IQR: 60.5-79.3 v.s. median: 70.6, IQR: 57.1-78.6, p=0.0136) after initial presentation of GAD diseases. In diastolic BP measurements, males had larger variance (median: 65.5, IQR: 30.3-81.0 v.s. median: 54.3, IQR: 31.4-88.9, p<0.0001) and larger variability score (median: 63.3, IQR: 40.2-71.2 v.s. median: 57.1, IQR: 46.2-66.7, p<0.0001) after initial presentation of GAD; females had larger minimal test (median: 63.0, IQR: 54.0-67.0 v.s. median: 58.0, IQR: 52.0-66.0, p=0.0003), larger variance (median: 61.8, IQR: 31.4-81.4 v.s. median: 56.1, IQR: 33.4-84.3, p<0.0001), and larger variability score (median: 62.3, IQR: 43.2-67.54 v.s. median: 55.8, IQR: 47.4-66.2, p<0.0001).

**Table 4.**
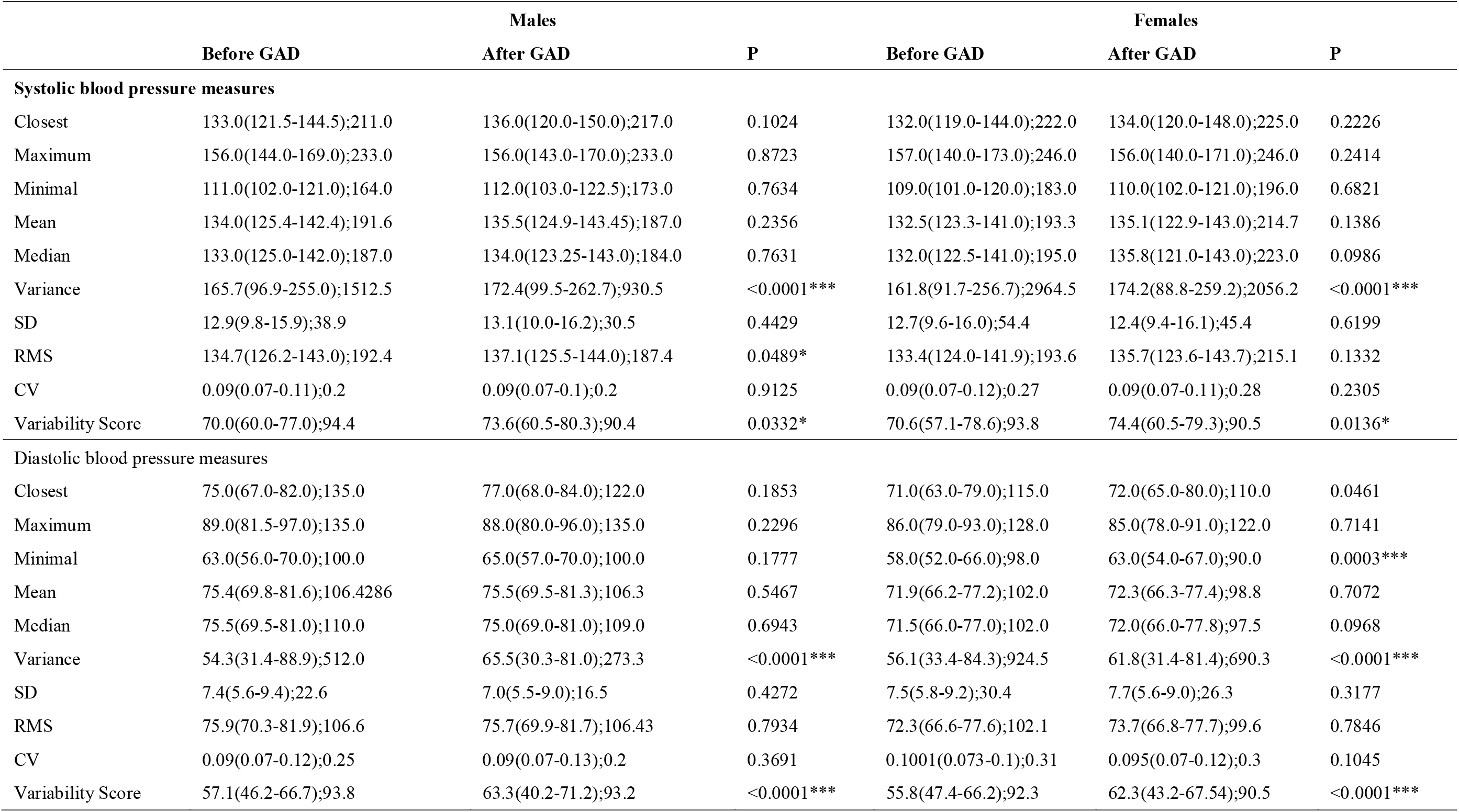
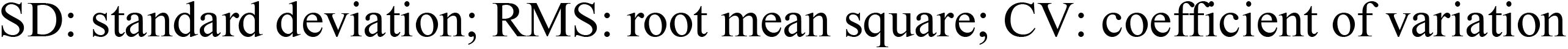
Gender-specific blood pressure measure changes before and after GAD development. * for p≤ 0.05, ** for p ≤ 0.01, *** for p ≤ 0.001

|  | Males |  |  | Females |  |  |
| --- | --- | --- | --- | --- | --- | --- |
|  | Before GAD | After GAD | P | Before GAD | After GAD | P |
| **Systolic blood pressure measures** |  |  |  |  |  |  |
| Closest | 133.0(121.5-144.5);211.0 | 136.0(120.0-150.0);217.0 | 0.1024 | 132.0(119.0-144.0);222.0 | 134.0(120.0-148.0);225.0 | 0.2226 |
| Maximum | 156.0(144.0-169.0);233.0 | 156.0(143.0-170.0);233.0 | 0.8723 | 157.0(140.0-173.0);246.0 | 156.0(140.0-171.0);246.0 | 0.2414 |
| Minimal | 111.0(102.0-121.0);164.0 | 112.0(103.0-122.5);173.0 | 0.7634 | 109.0(101.0-120.0);183.0 | 110.0(102.0-121.0);196.0 | 0.6821 |
| Mean | 134.0(125.4-142.4);191.6 | 135.5(124.9-143.45);187.0 | 0.2356 | 132.5(123.3-141.0);193.3 | 135.1(122.9-143.0);214.7 | 0.1386 |
| Median | 133.0(125.0-142.0);187.0 | 134.0(123.25-143.0);184.0 | 0.7631 | 132.0(122.5-141.0);195.0 | 135.8(121.0-143.0);223.0 | 0.0986 |
| Variance | 165.7(96.9-255.0);1512.5 | 172.4(99.5-262.7);930.5 | <0.0001*** | 161.8(91.7-256.7);2964.5 | 174.2(88.8-259.2);2056.2 | <0.0001*** |
| SD | 12.9(9.8-15.9);38.9 | 13.1(10.0-16.2);30.5 | 0.4429 | 12.7(9.6-16.0);54.4 | 12.4(9.4-16.1);45.4 | 0.6199 |
| RMS | 134.7(126.2-143.0);192.4 | 137.1(125.5-144.0);187.4 | 0.0489* | 133.4(124.0-141.9);193.6 | 135.7(123.6-143.7);215.1 | 0.1332 |
| CV | 0.09(0.07-0.11);0.2 | 0.09(0.07-0.1);0.2 | 0.9125 | 0.09(0.07-0.12);0.27 | 0.09(0.07-0.11);0.28 | 0.2305 |
| Variability Score | 70.0(60.0-77.0);94.4 | 73.6(60.5-80.3);90.4 | 0.0332* | 70.6(57.1-78.6);93.8 | 74.4(60.5-79.3);90.5 | 0.0136* |
| **Diastolic blood pressure measures** |  |  |  |  |  |  |
| Closest | 75.0(67.0-82.0);135.0 | 77.0(68.0-84.0);122.0 | 0.1853 | 71.0(63.0-79.0);115.0 | 72.0(65.0-80.0);110.0 | 0.0461 |
| Maximum | 89.0(81.5-97.0);135.0 | 88.0(80.0-96.0);135.0 | 0.2296 | 86.0(79.0-93.0);128.0 | 85.0(78.0-91.0);122.0 | 0.7141 |
| Minimal | 63.0(56.0-70.0);100.0 | 65.0(57.0-70.0);100.0 | 0.1777 | 58.0(52.0-66.0);98.0 | 63.0(54.0-67.0);90.0 | 0.0003*** |
| Mean | 75.4(69.8-81.6);106.4286 | 75.5(69.5-81.3);106.3 | 0.5467 | 71.9(66.2-77.2);102.0 | 72.3(66.3-77.4);98.8 | 0.7072 |
| Median | 75.5(69.5-81.0);110.0 | 75.0(69.0-81.0);109.0 | 0.6943 | 71.5(66.0-77.0);102.0 | 72.0(66.0-77.8);97.5 | 0.0968 |
| Variance | 54.3(31.4-88.9);512.0 | 65.5(30.3-81.0);273.3 | <0.0001*** | 56.1(33.4-84.3);924.5 | 61.8(31.4-81.4);690.3 | <0.0001*** |
| SD | 7.4(5.6-9.4);22.6 | 7.0(5.5-9.0);16.5 | 0.4272 | 7.5(5.8-9.2);30.4 | 7.7(5.6-9.0);26.3 | 0.3177 |
| RMS | 75.9(70.3-81.9);106.6 | 75.7(69.9-81.7);106.43 | 0.7934 | 72.3(66.6-77.6);102.1 | 73.7(66.8-77.7);99.6 | 0.7846 |
| CV | 0.09(0.07-0.12);0.25 | 0.09(0.07-0.13);0.2 | 0.3691 | 0.1001(0.073-0.1);0.31 | 0.095(0.07-0.12);0.3 | 0.1045 |
| Variability Score | 57.1(46.2-66.7);93.8 | 63.3(40.2-71.2);93.2 | <0.0001*** | 55.8(47.4-66.2);92.3 | 62.3(43.2-67.54);90.5 | <0.0001*** |

SD: standard deviation; RMS: root mean square; CV: coefficient of variation

## Discussion

The main findings of this study are that: 1) higher baseline, maximum, minimum, SD, CV, and variability score of diastolic BP significantly predicted GAD, as did all systolic BP measures (baseline, latest, maximum, minimum, mean, median, variance, SD, RMS, CV, variability score), and 2) female and older patients with higher blood pressure and higher BPV were at the greatest risks of GAD.

The effects of GAD on BP and as a potential risk factor have been extensively examined in previous studies (Yu Pan et al., 2015). However, whether BP influences the risk of incident GAD has not been investigated in detail, and mixed results were seen in observational studies. Individuals with hypertension may be more likely to develop anxiety (Y. Pan et al., 2015; Player & Peterson, 2011), but this association is seen only when hypertension coexists with another chronic condition (Grimsrud, Stein, Seedat, Williams, & Myer, 2009), or when the patients are aware of their hypertension diagnosis (Hamer, Batty, Stamatakis, & Kivimaki, 2010). Previously, higher beat-to-beat BPV has been associated with incident GAD (Virtanen et al., 2003). Longer term visit-to-visit BPV has also been reported as an independent predictor of neurological conditions such as cognitive impairment in cohort studies (Nagai et al., 2012; Sabayan et al., 2013; Yano et al., 2014), but whether it can do so for incident GAD has never been explored. In this population-based study of patients attending family medicine clinics in the public sector of Hong Kong, we established for the first time the predictive value of different metrics of BP and BPV on incident GAD.

Whilst the physiological mechanisms underlying the bidirectional relationship between hypertension and incident GAD remain unclear, the phenomenon was reported in recent studies. Population-based studies demonstrated that patients with baseline anxiety had an increased risk of essential hypertension. (Bacon, Campbell, Arsenault, & Lavoie, 2014; Perez-Pinar et al., 2016; Stein et al., 2014) By contrast, a territory-wide study of over two million patients in Sweden demonstrated that hypertensive patients were more likely to suffer from GAD. (Sandstrom, Ljunggren, Wandell, Wahlstrom, & Carlsson, 2016) The presence of anxiety increases the risk of poor drug compliance amongst hypertensive patients, thus worsens their BP control. (Bautista, Vera-Cala, Colombo, & Smith, 2012)

Various hypotheses have been proposed to explain the association between GAD and hypertension. First of all, chronic stress, which induces a persistent maladaptive stress response that develops into GAD, results in long term cortisol elevation. (Patriquin & Mathew, 2017) Consequently, the hypothalamic-pituitary-adrenal axis becomes dysregulated and leads to hypertension. (Gold et al., 2005) Furthermore, it is postulated that exaggerated neurobiological sensitivity towards threat results in prolonged activation of the hypothalamic-pituitary-adrenal axis, which results in both the autonomic dysregulation underlying hypertension, and the biological change under GAD. (O’Donovan, Slavich, Epel, & Neylan, 2013) Other mechanisms including increased oxidative stress, physical inactivity and hypercapnia were reported to be common in both the pathogenesis of hypertension and GAD, thus may contribute to the association between the two diseases. (Johnson et al., 2012; Salim et al., 2010; Ushakov, Ivanchenko, & Gagarina, 2016)

Similarly, hypotheses have been proposed to explain the predictive value of BPV for incident GAD. Increased BPV has been shown to be due to reduced baroreflex sensitivity, which may reflect sympathovagal imbalance likely due to sympathetic hyperactivity, which is observed in GAD patients. (Januzzi, Stern, Pasternak, & DeSanctis, 2000; Tully & Tzourio, 2017; Virtanen et al., 2003) The BP instability may reflect compensatory hemodynamic changes under reduced arterial compliance and increased aortic stiffness under systemic inflammatory response, which is both a cause of hypertension and a consequence of GAD. (Mitchell, 2009; Rothwell, 2010) Furthermore, the pathological worrying in GAD may be associated with increased compliance towards antihypertensives, which are known to increase BPV. (Parati, Ochoa, Lombardi, & Bilo, 2013; Parker, Hyett, Hadzi-Pavlovic, Brotchie, & Walsh, 2011) Moreover, the use of medications such as beta blockers also predicted incident GAD. It may be that anxious patients may more likely to receive such medications to reduce the symptoms of anxiety.

### Limitations

Several limitations should be noted for the present study. Given its observational nature, there is information bias with regards to issues of under-coding, missing data and documentation errors. Moreover, data on lifestyle risk factors of hypertension, such as smoking and alcoholism, were unavailable, thus their potential influence on the relationship between BP and GAD cannot be assessed. Furthermore, the clinical circumstances of BP measurement during hospital visits were susceptible to the effects of circumstantial factors, which may introduce additional variables that affect patients’ BP and BPV.

## Conclusions

The relationships between longer term visit-to-visit BPV and incident GAD were identified. Female and older patients with higher blood pressure and higher BPV were at the highest risks of GAD. Future studies should examine the interacting effects between BPV and medication use to influence incident GAD and GAD-related outcomes.

## Data Availability

Data available upon request.

## Acknowledgements

None.

## Competing or Conflicts of Interest

None.

## Author Contributions

JZ, XW: data analysis, data interpretation, statistical analysis, manuscript drafting, critical revision of manuscript

SL, WTW, WKKW, KSKL, TL: project planning, data acquisition, data interpretation, critical revision of manuscript

BMYC: study supervision, data interpretation, statistical analysis, critical revision of manuscript QZ, GT: study conception, study supervision, project planning, data interpretation, statistical analysis, manuscript drafting, critical revision of manuscript

