## Supplementary Appendix for "Gender- and Age-Specific Associations of Visit-To-Visit Blood Pressure Variability with Incident Generalized Anxiety Disorder"

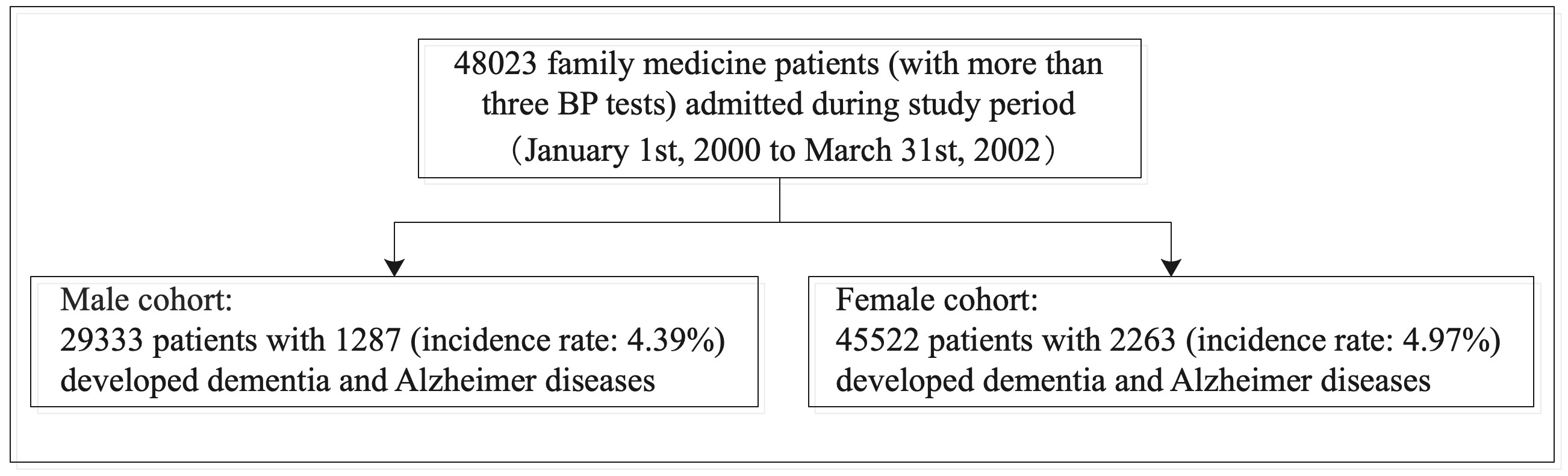

**Supplementary Figure 1. Flowchart of patient identification.**

**Supplementary Table 1. ICD-9 Codes for Comorbidities**

| **Comorbidity** | **Codes** |
| --- | --- |
| Cardiovascular | 428, 428.1, 428.2, 428.2, 428.21, 428.22, 428.23, 428.3, 428.3, 428.31, 428.32, 428.33, 428.4, 428.4, 428.41, 428.42, 428.43, 428.9 398.91, 402.01, 402.11, 402.91, 404.01, 404.03, 404.11, 404.13, 404.91, 404.93, 428 427.31, 429.4 |
| Respiratory | 490, 491, 492, 493, 494, 495, 496, 491.1, 491.2, 491.21, 491.22, 491.8, 491.9, 492.8, 493.01, 493.02, 493.1, 493.11, 493.12, 493.2, 493.21, 493.22, 493.8, 493.81, 493.82, 493.9, 493.91, 493.92, 494.1, 495.1, 495.2, 495.3, 495.4, 495.5, 495.6, 495.7, 495.8, 495.9 786.09, 518.81, 780.53, 137, E912, 465.9, 518.81, 518.81, 79.6, 518.81, 519.8, 780.59, 799.1, 780.57, 518.82, 480.1, 786.3, 519.8, 997.3, 165.9, 519.9, 648.91, 162.9, 162.3, 197, 162.5, 162.4, 486, 518.89, 496, 162.8, 415.1, 518, 162.9, 11.96, 482.1, 507, 513, 11.9, 511.8, 511.1, 11.94, 516.8, 793.1, 482.4, 507, 515, 197, 11.93, 482, 482.83, 518.4, 482.3, 482.2, 415.1, 502, 518.89, 235.7, 793.1, 934.8, 516.9, 136.3, 38.49, 506, 112.4, 487, 481, 117.9, 38.2, 518, 11.95, 79.89, 518.1, 480.9, 505, 516.8, 495.9, 518.3, 11.23, 416.8, 513, 397.1, 117.3, 483, 508, 998.81, 416, 514, 861.21, 502, 934.8, 480.8, 648.93, 11.2, 492.8, 484.6, 78.5, 484.1, 516.3, 415.1, 416.9, 415, 416.9, 429.89, 415, 747.49, 745, 417, 770.7, 427.5, 416.9, 416, 416.8, 746.02, 573.8, 642.9, 416, 747.3, 747.3, 770.3, 779.8, 515, 424.3, 416, 417.8, 747.3, 747.3, 745.4, 518.81, 786.09, V12.6, 478, 748.5, 162.9, 996.84, 748.5, 748.6, V42.1, 748.5, 11.05, 162, 518, 747.42, 518.89, 748.5, 517.2 |
| Renal | 198.7, 189, 189, 585.9, V56.0, 189.1, 584.9, 189.1, 593.9, 189, 189, 189.8, 239.5, 189, 583.81, 593.9, 591, V10.52, 250.4, 591, 255.4, 590.8, 586, 585.1, 591, 189, 591, 239.7, 788, 996.39, 189.1, 588.9, 592, 996.39, 250.4, 593.2, 255.4, 572.4, 194, 255, 453.3, 198, 585.9, 996.39, 996.39, 590.1, 591, 591, 223, 591, 584.9, 585.9, 250.41, 996.39, 581.9, 227, 593.5, 583.89, 593.89, 404.93, 255.5, 788.9, 250.43, 227, 405.92, 592, 753.12, 996.39, E879.1, V42.0, 592, 580.89, 403.9, 593.9, V59.4, 585.9, 580.9, 250.41, 996.39, 585.9, 794.4, 584.8, 584.5, 255.9, 441.4, V58.49, 404.11, 593.9, 592, 585, 759.1, 753.11, E879.1, 753.15, 585.9, 274, 588.8, 403.91, 404.9, 404.91, 404.92, 403, 404.01, 779.8, 250.4, 227, 227, 227, 404.93, 996.81, 593.89, 753.8, 593.2, 592, 753, 996.81, 223, 589.1, 996.81, 582.9, 585.9, 996.81, 753.1, 753.3, E878.0, 593.9, 753.3, 753.17, 583.9, 593.9, 589, 866 |
| Endocrine | 202.8, 200.1, 200.12, 201.9, 204, 202.88, 200.18, 196, 204.01, 785.6, 200.11, 200.13, 202.8, 785.6, 202.85, 202.81, 202.8, 202.8, 196, 196.9, 202.8, 202.82, 785.6, 202.8, 202, 202.87, 785.6, 202.84, 202.01, 196.8, 457.1, 12.1, 785.6, V10.79, 196.5, 196.2, 238.7, 196.1, 200.14, 457.2, 238.7, V10.61, 201.9, 457, 202.8, 289.3, 245.2, 238.7, 785.6, 457.9, 785.6, 202.93, 196.9, 202.97, 757, V10.71, 288.8, 204.1, 202.83, 457.1, 289.3, 785.6, V77.9, 237.4, 239.7, 198.89, 623.5, 259.9, 200.2, V10.71 |
| Diabetes mellitus | 251.2, 362.01, 362.02, 250.4, 250.82, 790.2, 790.6, 250.5, 250.5, 250.6, 357.2, 790.2, 250, 250.4, 250.6, 250.8, 250.51, 250.5, 250.8, 250.82, 250.51, 250.51, 250.12, 251.2, 250.12, 250.5, 250.83, 251.2, 250.41, 251.1, 250.52, 250.5, 648.81, 250.43, 250.53, 250.53, 250.81, 250.22, 250.13, 250.22, 250.83, 250.41, 250.5, 250.52, 250.52, 250.82, 253.5, V18.0, 588.1 |
| Hypertension | 401.9, 401.9, 250.82, 790.6, 401.9, 401.9, 250.82, 796.2, 402.9, 250.83, 405.99, 642.93, 642.01, 642.91, 401.9, E942.6, 405.09, 403.9, 437.2, 401, 401.1, 401, 642.33, 348.2, 779.8, 365.04, 572.3, 416, 416, 405.91, 416.8, 642.3 |
| Gastrointestinal | 153.3, 154.1, 153.9, 569.89, 154, 153.1, 578.9, 560.9, 569.3, 537.89, 558.9, 562.1, 153.6, 239, 532.3, 569.89, 532.7, 535.6, 558.9, 38.42, 569.89, 8.45, 153.2, 569.49, 79.89, 532.9, 569, 154.1, 41.4, 537.89, 152.1, 578.9, 787.8, 197.4, 535.5, 9, 569.83, 569.6, 153.4, 560.9, 537.3, 41.04, 569.84, 239, 569.81, 8.8, 535, 560.9, 532, 532.4, 560.81, 235.2, 38.49, 8.45, 235.2, 532.9, 569.81, 537.89, 557.9, 569.41, 997.4, 14.8, 787.99, 8.46, 535.5, 569.41, 997.4, 578.9, 569.82, 537.9, 560.1, 569.82, 557.9, 211.3, 556.9, 562, 558.9, 578.9, 536.9, 8.46, 535.6, 566, 569.49, 564.3, 569.89, 564.8, 8.46, 569.83, 997.4, 997.4, 997.4, 562.11, 211.2, 9.1, 211.3, 8.47, 8.5, 211.3, 569.83, 532.1, 535.61, 560, 569.83, 565.1, 619.1, 152.9, 568, 566, 569.43, 152, 8.46, 562.11, 8.61, 569.83, 569.83, 569.81, 596.1, 535.5, 151.4, 151.9, 151.5, 151.8, 151.1, 456.8, 531.7, 535.4, 531.3, 537.89, 211.1, 531.9, 235.2, 531, 531.4, 456.8, 535.1, 151.3, 230.2, 151.6, 535, 211.1, 536.3, 535, 535.51, 578.9, 531.1, 535.1, 456.8, 531.5, 537.84, 535.01, 530.7, 535.1, 535.2, 535.5, 537.6, 202.83, 535.1, 531.4, 558.9, 558.9, 558, 569.85, 153, 555.1, 562.1, 562.13, 562.11, 562.12, 569.83, 560.2, 230.4, 569.3, 154 |
| Stroke | 435, 435.1, 435.2, 435.3, 435.8, 435.9, 433.81, 433.91, 434, 436, 437, 437.1, 433.31, 433.01, 434.01, 434.1, 434.11, 434.9, 434.91, 437.2, 437.3, 437.4, 437.5, 437.6, 437.7, 437.8, 437.9, |

**Supplementary Table 2. Blood pressure characteristics of patients developing incident GAD in different age-specific groups**

* for p≤ 0.05, ** for p ≤ 0.01, *** for p ≤ 0.001

|  | **[10,20) (N=3)** | **[20,30) (N=25)** | **[30,40) (N=113)** | **[40,50) (N=431)** | **[50,60) (N=514)** | **[60,70) (N=588)** | **[70,80) (N=418)** | **[80,90) (N=81)** |
| --- | --- | --- | --- | --- | --- | --- | --- | --- |
|  | **Median (IQR); Max** | **Median (IQR); Max** | **Median (IQR); Max** | **Median (IQR); Max** | **Median (IQR); Max** | **Median (IQR); Max** | **Median (IQR); Max** | **Median (IQR); Max** |
| **Diastolic blood pressure measures** | | | | | | | | |
| Baseline, mm Hg | 66.0(62.5-68.0);70.0 | 69.0(65.0-74.0);94.0 | 70.0(60.0-80.0);110.0 | 73.0(65.0-81.5);122.0 | 75.0(66.0-82.0);110.0 | 73.0(66.5-80.0);118.0 | 72.0(65.0-80.0);110.0 | 71.0(61.0-80.0);110.0 |
| Latest, mm Hg | 60.0(57.0-63.0);66.0 | 70.0(64.0-81.0);95.0 | 74.0(64.0-80.0);114.0 | 74.0(67.0-80.0);121.0 | 72.0(65.0-79.0);113.0 | 71.0(64.0-78.0);135.0 | 69.0(60.5-76.0);115.0 | 69.0(59.0-76.0);90.0 |
| Maximum, mm Hg | 74.0(72.0-83.5);93.0 | 80.0(72.0-87.0);112.0 | 81.0(72.0-92.0);118.0 | 88.0(78.0-96.0);122.0 | 88.0(80.0-95.0);122.0 | 88.0(81.0-95.0);135.0 | 85.0(79.0-90.0);127.0 | 83.0(78.0-90.0);110.0 |
| Minimal, mm Hg | 60.0(57.0-63.0);66.0 | 65.0(58.0-70.0);77.0 | 60.0(55.0-68.0);95.0 | 61.0(55.0-70.0);100.0 | 60.0(53.0-69.0);92.0 | 58.0(52.0-65.0);88.0 | 57.0(50.0-64.0);90.0 | 54.0(50.0-64.0);90.0 |
| Mean, mm Hg | 65.0(64.25-70.1);75.2 | 69.6721(66.5-78.0);89.5 | 72.9024(63.75-80.0);106.4 | 74.63(68.52-80.51);106.3 | 74.7(68.75-79.0);97.0 | 72.3812(67.5-77.0625);93.62 | 70.0469(65.3694-74.5);98.8 | 69.3333(64.0625-74.0);90.0 |
| Median, mm Hg | 65.0(64.0-70.0);75.0 | 69.0(65.5-78.0);90.0 | 74.0(63.0-80.0);110.0 | 74.0(68.0-80.0);109.0 | 74.0(69.0-79.5);101.0 | 72.0(67.5-77.0);93.0 | 69.25(65.0-75.0);97.5 | 70.0(64.0-75.0);90.0 |
| Variance | 77.7(63.8-99.7);121.7 | 32.0(23.0-74.25);258.3 | 50.0(24.5-81.0);412.3 | 47.8(28.3-73.1);512.0 | 51.4(31.0-73.7);331.2 | 61.1(37.9-89.5);612.5 | 62.5(38.6-93.8);690.3 | 82.8(42.9-115.65);924.5 |
| SD | 8.81(7.9-9.92);11.03 | 5.66(4.8-8.62);16.1 | 7.07(4.95-9.0);20.31 | 6.91(5.32-8.55);22.63 | 7.17(5.57-8.58);18.2 | 7.82(6.15-9.46);24.75 | 7.92(6.2-9.7);26.3 | 9.1(6.6-10.8);30.4 |
| RMS | 65.2(64.6-70.5);75.8 | 69.9(67.7-78.1);89.9 | 73.3(64.1-80.2);106.6 | 74.98(68.9-80.8);106.4 | 74.79(69.18-79.43);97.2 | 72.7(68.1-77.5);94.8 | 70.6(65.9-74.8);99.5 | 69.52(65.22-74.71);91.1 |
| CV | 0.12(0.099-0.13);0.13 | 0.07(0.05-0.11);0.21 | 0.09(0.06-0.11);0.26 | 0.09(0.06-0.112);0.22 | 0.09(0.07-0.114);0.21 | 0.11(0.08-0.13);0.25 | 0.11(0.08-0.13);0.31 | 0.13(0.09-0.15);0.24 |
| Variability score | 60.0(55.0-67.5);75.0 | 50.0(33.3-55.7);87.5 | 50.0(33.33-60.0);88.89 | 54.6(42.9-64.3);93.75 | 54.55(45.58-63.64);90.91 | 58.68(50.0-66.7);90.0 | 59.12(50.0-66.7);90.91 | 59.5238(50.0-70.8);92.0 |
| **Systolic blood pressure measures** | | | | | | | | |
| Baseline, mm Hg | 124.0(120.5-124.0);124.0 | 110.0(106.0-118.0);154.0 | 114.0(105.0-126.0);192.0 | 123.0(110.5-139.0);205.0 | 130.0(116.0-144.0);208.0 | 138.0(123.0-151.0);218.0 | 141.0(130.0-156.0);225.0 | 143.0(133.0-161.0);210.0 |
| Latest, mm Hg | 101.0(97.5-117.0);133.0 | 120.0(104.0-128.0);154.0 | 121.0(108.0-133.0);175.0 | 126.0(115.0-137.0);208.0 | 130.0(120.0-139.0);200.0 | 135.0(123.0-145.0);218.0 | 136.0(123.0-149.0);234.0 | 136.0(123.0-156.0);187.0 |
| Maximum, mm Hg | 124.0(124.0-140.0);156.0 | 128.0(116.0-146.0);184.0 | 133.0(119.0-150.0);192.0 | 147.0(130.0-160.0);213.0 | 155.0(142.0-167.0);235.0 | 163.0(150.0-178.5);246.0 | 163.0(151.0-178.5);234.0 | 170.0(157.0-187.0);230.0 |
| Minimal, mm Hg | 100.0(97.0-102.0);104.0 | 103.0(99.0-110.0);133.0 | 104.0(96.0-111.0);150.0 | 108.0(100.0-115.0);159.0 | 109.0(101.0-118.0);156.0 | 110.0(102.0-121.0);183.0 | 113.0(104.0-123.0);177.0 | 123.0(106.0-132.0);163.0 |
| Mean, mm Hg | 110.3(109.63-118.43);126.6 | 114.0(108.5-128.6481);147.44 | 116.7(108.0-129.33);160.7 | 126.5(116.9-134.7);173.0 | 131.6(123.7-138.2);179.4918 | 135.9(128.3-144.4);191.6 | 138.3(130.5-145.9);193.3 | 144.1(135.4-152.3);172.0 |
| Median, mm Hg | 109.0(108.75-116.0);123.0 | 113.5(109.0-130.0);149.0 | 118.5(107.0-130.0);160.0 | 126.0(116.0-134.0);171.0 | 131.0(123.0-138.0);180.0 | 136.0(127.5-144.0);195.0 | 138.5(130.0-146.0);192.5 | 143.0(134.0-154.0);172.0 |
| Variance | 380.3(258.9416-415.15);450.0 | 82.8(33.3-128.0);263.6 | 98.0(50.0-166.5636);512.0 | 117.6(64.1-194.0);930.5 | 145.8(91.7-206.3);846.5 | 188.7(117.7-280.4);1584.7 | 220.7(139.2-330.2);1512.5 | 251.5(144.98-420.8);2964.5 |
| SD | 19.5(15.6-20.4);21.2 | 9.1(5.8-11.3);16.2 | 9.9(7.1-12.91);22.63 | 10.85(8.01-13.93);30.5 | 12.1(9.6-14.4);29.1 | 13.7(10.9-16.7);39.81 | 14.9(11.8-18.2);38.9 | 15.9(12.0-20.5);54.45 |
| RMS | 110.72(110.4-119.3);127.8 | 114.3(108.6-129.04);147.7 | 117.5(108.5-130.2);160.8 | 126.97(117.2-135.1);173.5 | 132.4(124.3-138.8);180.5 | 136.5(129.0-144.9);192.4 | 138.98(131.2-146.7);193.6 | 145.1(136.4-152.9);173.9 |
| CV | 0.1376(0.1149-0.1377);0.14 | 0.06(0.05-0.09);0.12 | 0.07(0.05-0.1);0.15 | 0.08(0.06-0.1);0.2 | 0.09(0.07-0.11);0.2 | 0.097(0.08-0.12);0.27 | 0.10(0.08-0.13);0.24 | 0.1(0.07-0.14);0.24 |
| Variability score | 50.0(50.0-65.0);80.0 | 50.0(50.0-66.7);87.5 | 64.3(50.0-72.3);85.7 | 66.7(50.0-75.0);93.75 | 70.14(60.0-76.61);94.4 | 73.7(63.1-80.0);93.75 | 74.60(61.33-80.0);93.3 | 71.43(50.0-81.25);92.86 |
